# Safety and Efficacy of Adjunctive Intra-arterial Tenecteplase following Successful Thrombectomy in Patients with Large Vessel Occlusion: Study Rationale and Design of A Phase 1/2 Randomized Clinical Trial

**DOI:** 10.1101/2024.11.18.24317521

**Authors:** Xianhua Hou, Jiacheng Huang, Changwei Guo, Li Wang, Yuxuan He, Lin Chen, Qu Liu, Junhua Wu, Min Wu, Yazhou Wu, Thanh N. Nguyen, Raul G. Nogueira, Duolao Wang, Jeffrey L. Saver, Wenjie Zi, Zhenhua Zhou

**Affiliations:** Department of Neurology, Southwest Hospital, Army Medical University, Chongqing, China; Department of Neurology, Xinqiao Hospital, Army Medical University, Chongqing, China; Department of Neurology, The Second Affiliated Hospital of Chongqing Medical University, Chongqing, China; Department of Neurology, Zigong Third People’s Hospital, Zigong, Sichuan, China; Department of Health Statistics, College of Preventive Medicine, Army Medical University, Chongqing, China; Department of Neurology and Radiology, Boston Medical Center, USA; UPMC Stroke Institute, Departments of Neurology and Neurosurgery, University of Pittsburgh School of Medicine, PA, USA; Global Health Trials Unit, Liverpool School of Tropical Medicine, Liverpool, UK; Department of Neurology and Comprehensive Stroke Center, David Geffen School of Medicine, University of California, Los Angeles, CA, USA

**Keywords:** Clinical trial, Tenecteplase, Mechanical thrombectomy, Acute ischemic stroke, Large vessel occlusion

## Abstract

**Background:** Adjunct intra-arterial thrombolysis has been shown to potentially improve clinical outcomes in patients with large vessel occlusion (LVO) stroke who have undergone successful endovascular thrombectomy (EVT). Tenecteplase, known for its enhanced fibrin specificity and extended activity duration, may be a better choice than alteplase for intra-arterial thrombolysis. However, the optimal dose, safety and efficacy of intra-arterial tenecteplase remain unclear.

**Objective:** To evaluate the optimal dose, safety and efficacy of adjunctive intra-arterial tenecteplase following successful EVT in LVO patients.

**Methods and design:** This study is a two-part, phase Ib/IIa, multicenter, open-label, 14+8 dose-escalation (Part I) and dose-expansion (Part II) trial conducted in China involving patients LVO patients who achieved successful EVT (defined as the expanded Thrombolysis in Cerebral Infarction [eTICI] grade of 2b50 or higher) within 24 hours of last known well. In Part Ib, the dose escalation will be conducted in up to four tiers of 14+8 patients, starting at 0.03125 mg/kg, to a planned maximum of 0.1875mg/kg, and the primary outcome is symptomatic intracranial hemorrhage within 24 hours. In Part IIa, 157 patients will be randomized among two selected safe doses and a placebo, and the primary outcome will be the proportion of patients with nondisabled outcome (modified Rankin Scale score of 0 to 1) at 90 days.

**Discussions:** This pivotal trial will provide important data on adjunctive intra-arterial tenecteplase following successful EVT in patients with acute ischemic stroke due to LVO and may support advance of treatment standards.

**Trial registry number:** ChiCTR2300073787 and ChiCTR2400080624 (www.chictr.org.cn).

## Introduction

Stroke is one of the leading causes of morbidity and mortality worldwide.^1^ Endovascular thrombectomy (EVT) is currently recommended as the gold standard treatment for acute ischemic stroke caused by large vessel occlusion (LVO).^2^ However, a meta-analysis study consisted of five randomized clinical trials indicated that two-thirds of patients had disabled (modified Rankin Scale [mRS] 2-6) outcomes after EVT despite successful reperfusion (defined as the extended Thrombolysis In Cerebral Infarction [eTICI] grade of 2b50 or higher.^3,4^ Already infarcted tissue present before the procedure, combined with infarct expansion into areas lacking sufficient macro- and microcirculatory reperfusion afterward, may lead to incomplete functional recovery.^5^ Previous studies demonstrated that even when successful reperfusion has been achieved after EVT, hypoperfusion is common and this is associated with poor clinical outcomes.^6^ Preclinical studies have shown that capillary occlusion, perivascular space obstruction, or distal microembolism from a proximal thrombus may cause incomplete reperfusion of the microcirculation and distal vasculature.^7^

Targeting the mirco-thrombi, the Effect of Intra-arterial Alteplase vs Placebo Following Successful Thrombectomy on Functional Outcomes in Patients With Large Vessel Occlusion Acute Ischemic Stroke (CHOICE) study showed that adjuvant intra-arterial thrombolysis after successful reperfusion by EVT could improve excellent functional outcome at 90 days.^8^ However, this study was terminated prematurely with a resulting limited sample size.

Tenecteplase is a modified form of alteplase, with a longer half-life and greater fibrin specificity.^9,10^ After extensive exploration, preliminary research established the optimal dose of intravenous TNK (0.25 mg/kg) for thrombolysis therapy in stroke.^11^ In the Tenecteplase versus Alteplase before Thrombectomy for Ischemic Stroke (EXTEND-IA TNK) study, administering tenecteplase (0.25 mg/kg) prior to thrombectomy resulted in a higher rate of reperfusion and improved functional outcomes compared to alteplase (0.9 mg/kg) in patients with LVO stroke.^10,12^ Considering the advantages of tenecteplase, adjuvant intra-arterial administration of tenecteplase may provide greater potential benefits for patients with LVO stroke following successful reperfusion compared with alteplase. However, the optimal dose, efficacy, and safety of intra-arterial tenecteplase following successful reperfusion is unknown.

Therefore, we conduct a two-part, phase Ib/IIa, multicenter, open-label, 14+8 dose-escalation (Part 1) and dose-expansion (Part 2) trial to determine the most promising dose to advance to a pivotal trial of intra-arterial tenecteplase following successful thrombectomy in patients with acute ischemic stroke due to LVO within 24h of symptom onset.

## Methods

### Study design

The Safety and Efficacy of Adjunctive Intra-arterial Tenecteplase following Successful Thrombectomy in Patients with Large Vessel Occlusion (DATE) is an open-label, randomized, multicenter, Phase Ib/IIa clinical trial designed to identify a dose of intra-arterial tenecteplase following successful EVT in patients with acute ischemic stroke due to LVO that shows sufficient evidence of safety and efficacy to advance to a pivotal trial. In designing this protocol, we adhered to the SPIRIT reporting guidelines^13^ for clarity and consistency. The DATE trial will be divided into two parts: the first part (Phase Ib) is a pilot dose-escalation safety study, and the second part (Phase IIa) is an exploratory study to assess the safety and potential efficacy of adjunctive intra-arterial tenecteplase after successful EVT in patients with LVO stroke at two different doses. After the completion of Phase Ib, a data safety monitoring board (DSMB) unblinded to study groups will select 2 doses (A or B) to be tested in Phase IIa according to the initial safety results.

#### Dose Escalation (Phase Ib)

This is a dose escalation trial. During this phase, participants will receive intra-arterial administration of increasing doses of tenecteplase after successful EVT. Patients will be treated with up to 4 escalating dose levels: 0.03125 mg/kg (1/8 *i*.*v* dose), 0.0625 mg/kg (1/4 *i*.*v* dose),0.125 mg/kg (1/2 *i*.*v* dose), and 0.1875 mg/kg (3/4 *i*.*v* dose).

According to the Chinese Acute Anterior Circulatory Ischemic Stroke Endovascular Treatment Registry Study, 13.8% of patients experienced symptomatic intracranial hemorrhage (sICH) within 24 hours of receiving EVT.^14^ Using sample sizes typical for dose escalation phase I clinical trials^15,16^, we will use a 14+8 design scheme with sICH occurrence within 24 hours after EVT as the dose-limiting toxicity. At each tier, 14 patients will initially be enrolled. If fewer than 2 of 14 subjects develop sICH, the trial will advance to the next tier. If 2 of the 14 develop sICH, 8 more patients will be enrolled at that dose. If 0-1 of the additional 8 patients develops sICH, the trial will advance to the next tier. If 2 of the additional 8 patients develops sICH or 3 of the first 14 develop sICH, that dose will be deemed not tolerated and the immediately preceding dose will be the estimated maximum tolerated dose. The chart is shown in Figure 1.

**Figure 1.**
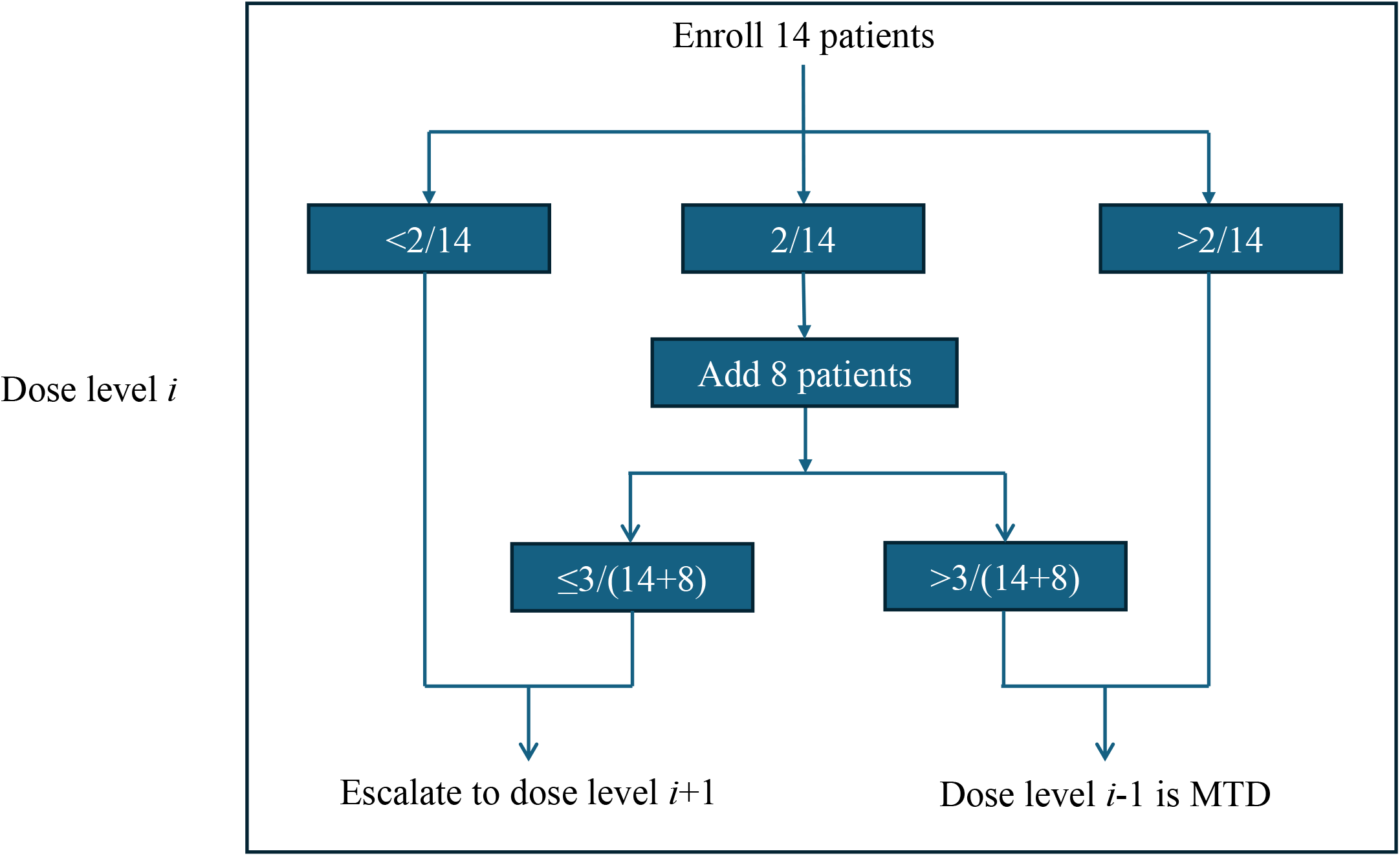
Flow chart for the Phase I of the DATE trial.

#### Dose Expansion (Phase IIa)

Two doses (A and B) will be chosen by the DSMB and investigators jointly according to the results in Phase Ib. In Phase II, a total of 157 new patients will be enrolled, with 46 patients assigned to the dose A group, 46 patients assigned to the dose B group, and an additional 65 patients allocated to the control group.

During this phase, 3 arms will be studied and patients will be randomized with stratification allocation according to two strata: patient age (<70 vs ≥ 70 years), and admission NIHSS score (< 15 vs ≥ 15), to receive 1 of the 2 selected doses of tenecteplase, or to the control group in a 1:1:√2 ratio, which in turn yields probabilities of assignment of 0.293,0.293, and 0.414, respectively. Patients will be randomly assigned by using a real-time internet-based system. This process is automated which allows for concealment of the sequence of allocation.

### Objectives

The primary objective of this study is to preliminarily assess the safety and efficacy of adjunctive intra-arterial tenecteplase at different doses following successful EVT in patients with acute ischemic stroke due to LVO to determine the promising dose to advance to a pivotal trial.

### Outcome

The study is designed to have two parts with different outcomes.

### Dose Escalation (Phase Ib)

#### Primary outcome

The primary outcome is sICH within 24 hours.

#### Secondary outcome

1. 90-day modified mRS score 0-1 (%);
2. Proportion of patients with functional independence (mRS score 0 to 2) at 90 days;
3. Level of disability (ordinal distribution of mRS scores) at 90 days;
4. Favorable shift in reperfusion on the eTICI score after intra-arterial tenecteplase thrombolysis therapy. All post-EVT, pre-IA and final post-IA final, angiograms will be scored at a core lab by central and blinded reviewers using the eTICI and classified as eTICI2b, eTICI2c, and eTICI3. The post treatment angiographies will be scored using the eTICI and classified as “improved”, “worsened” or “unchanged” with regard to the pre-IA infusion eTICI score.
5. The change from baseline of the NIHSS score to 5–7 days or discharge if earlier;
6. EQ-5D-3L at 90 days;
7. Mortality within 90 days.

### Dose Expansion (Phase IIa)

#### Primary outcome

The primary outcome will be the proportion of patients with a nondisabled outcome (mRS score of 0 or 1) at 90 (±14) days. Primary outcome assessments will be performed at 90 (±14) days by two independent, certified physicians who are blinded to the treatment details. To maintain the reliability, accessibility, and traceability of the mRS score, we will retain a video or audio records of the 90-day follow-up for all patients. If video or audio recordings are unavailable, outcomes will be determined in-person by certified local investigators, who will be also unaware of the treatment assignment.

#### Secondary outcomes

1. Proportion of patients with functional independence (mRS score 0 to 2) at 90 days;
2. Level of disability (ordinal distribution of mRS scores) at 90 days;
3. Favorable shift in eTICI score after intra-arterial tenecteplase thrombolysis therapy;
4. The change of the NIHSS score from baseline to 5–7 days or discharge;
5. EQ-5D-3L at 90 days;
6. sICH rate within 24 hours;
7. Mortality within 90 days.

### Inclusion and exclusion criteria

#### Inclusion criteria

1. Age ≥18 years;
2. Time from last known well within 24 hours;
3. Large vessel occlusive stroke in the anterior circulation confirmed by computed tomography angiography/magnetic resonance angiography, including intracranial segment of the internal carotid artery, first or second segment of the middle cerebral artery;
4. Alberta Stroke Program Early CT Score (ASPECTS) ≥ 6 based on non-contrast computed tomography;
5. National Institutes of Health Stroke Scale (NIHSS) score ≥ 6;
6. Successful endovascular thrombectomy (eTICI 2b-3);
7. Total pass numbers of thrombectomy procedure ≤ 3;
8. Written informed consent signed by patient or their family member.

#### Exclusion criteria

1. NIHSS score ≥ 25;
2. Intracranial hemorrhage confirmed by computed tomography or magnetic resonance imaging;
3. Treated by intravenous thrombolysis;
4. Prestroke mRS score ≥ 2.
5. Intraprocedural digital subtraction angiography suggests vessel penetration, dissection, or extravasation of contrast medium;
6. Pregnant or lactating patients;
7. Allergic to contrast agent or Tenecteplase;
8. Systolic pressure greater than 185 mmHg or diastolic pressure greater than 110 mmHg despite blood pressure lowering treatment;
9. Genetic or acquired bleeding disposition with anticoagulation factor deficiency or already taking oral anticoagulants within 48 hours and INR > 1.7;
10. Blood glucose < 2.8 mmol/L (50 mg/dl) or > 22.2 mmol/L (400 mg/dl), platelets < 90*10^9/L;
11. History of bleeding in the last 1 month (gastrointestinal and urinary tract bleeding);
12. Patients on chronic hemodialysis and severe renal insufficiency (glomerular filtration rate < 30 ml/min or blood creatinine > 220 μmol/L [2.5mg/dl]);
13. Any terminal illness with a life expectancy of less than 6 months;
14. Intracranial aneurysm, arteriovenous malformation;
15. Brain tumor with occupying mass effect on imaging;
16. Puncture to reperfusion time > 90 min;
17. Current participation in another clinical trial;
18. Unlikely to be available for 90-day follow-up.

### Participating center eligibility

To qualify for participation in this trial, study centers will be mandated to have performed a minimum of 50 endovascular procedures each year. Additionally, all neurointerventionists involved will be required to have over five years of experience in cerebrovascular interventions, along with a track record of having performed at least 30 cases of mechanical thrombectomy using stent retriever devices annually.

### Enrollment

Before initiating any procedure, participants or their legally authorized representatives will be provided with comprehensive oral and written information regarding the study, encompassing its objectives, potential risks, and anticipated benefits. Subsequently, they will be required to give their informed consent.

All individuals who fulfill the specified eligibility criteria are eligible for enrollment in the DATE clinical trial.

### Study Intervention

Eligible patients assigned to the tenecteplase group will undergo an infusion of intra-arterial tenecteplase with the assigned dose. This infusion will be administered through a distal access catheter or microcatheter positioned proximal to the initially occluded artery. Patients allocated to the control group will terminate the procedure without further intraarterial thrombolysis.

All enrolled patients should be monitored in the acute stroke unit and can be admitted to the intensive care unit if necessary. All enrolled patients will undergo standardized medical treatment management and subsequent secondary preventive medication according to the Chinese Guidelines for Endovascular Treatment of Acute ischemic Stroke 2018.^17^

### Blinding and masking

Each participating site will allocate one or more physicians to perform follow-up evaluations at 24-hour intervals, between five to seven days post-treatment, or at the time of discharge if it occurs sooner, along with a 90-day assessment. It is imperative that these physicians are not involved in the subject’s initial treatment to ensure their blinding to the treatment assignment.

### Data safety monitoring board

The independent Data and Safety Monitoring Committee (DSMB) is responsible for reviewing the aggregated data and determining the study safety endpoints. The DSMB regularly reviews the study data added during the trial and advises the sponsor on the continued safety of the subjects and those not yet included in the study. The DSMB advises on the continued effectiveness and scientific value of the study.

### Sample size estimates

#### Dose Escalation (Phase Ib)

The Endovascular Treatment for Acute Anterior Circulation Ischemic Stroke registry in China showed that the incidence of sICH within 24 hours after EVT was 13.8%.^14^ Based on this data, we designed a 14+8 enrollment plan as follows:

1. 14 subjects will be included in the trial for the first time for each dose;
2. If<2 DLT (dose limit toxicity) symptomatic intracranial hemorrhages occur within 24 hours after EVT, the trial advances to the next dose level;
3. If 2/14 DLT, then include 8 more people at the same dose;
4. If 2/14+0-1/8DLT (9.1% to 13.6%), proceed to the next dose level;
5. If 2/14+2/8 DLT (18.2%) or 3/14 DLT, then this dose is deemed unsafe and the prior lower dose is the MTD.

### Dose Expansion (Phase IIa)

In Phase IIa randomization will allocate to randomize 46 subjects each into dose group A and dose group B, and 65 subjects into the control group in a 1:1:√2 ratio. This approach is similar to the sample size calculation method used in the APRIL study.^18^

### Statistical analyses

Continuous variables will be summarized with the median, the inter-quartile range, the minimum, and the maximum. Categorical variables will be summarized with counts and percentages. Patient compliance with eligibility criteria and treatment administration, major protocol non-compliance, patient withdrawal and the reason for withdrawal (e.g., adverse event, protocol non-compliance, lost to follow up, failed to return, consent withdrawal, and other reasons) and assignment to each analysis population will be summarized using appropriate statistics by treatment arms.

The statistical results of outcome analyses will be provided with point estimates of treatment differences with 95% confidence intervals (CIs).

The primary outcome in both phase Ib and IIa will be analyzed using generalized linear models from which a risk ratio with its 95% CI will be estimated as a measurement of treatment effect. The secondary outcomes and safety outcomes (binary, continuous and ordinal) will be analyzed using generalized linear models. For non-normal continuous outcome, a win ratio will be estimated as a measurement of treatment effect. A pooled analysis that combines the phase Ib and phase IIa will be performed as supportive analysis. The primary data analyses will adhere to the intention-to-treat principle. Additionally, per-protocol analyses will be conducted as supplementary analyses.

All statistical analyses will be described in detail in the statistical analysis plan which will be finalized before the study database lock, and will be performed using SAS version 9.4 (SAS Institute) and R version 4.4.0 or higher (R Foundation for Statistical Computing). The reporting of trial results will conform to the Consolidated Standards of Reporting Trials guidelines for randomized trials.

### Ethical considerations

The study adheres to the ethical guidelines outlined in the Declaration of Helsinki. The protocol was approved by the Ethics Committee of the First Affiliated Hospital of the Army Medical University and all participating centers. Any modifications to the trial protocol will only be executed after obtaining further approval from the ethics committee.

## Discussion

Despite advancements in thrombectomy devices and workflow processes, outcomes for patients with LVO remain suboptimal. Adjunctive intra-arterial thrombolysis following successful reperfusion in patients with LVO stroke has the potential to improve functional outcomes, while the optimal dose remains unclear. To our knowledge, the DATE trial is the first to evaluate the safety, efficacy, and dose-response effects of intra-arterial tenecteplase in patients with LVO stroke after successful EVT.

The CHOICE study demonstrated the potential benefits of intra-arterial alteplase at a dose equivalent to 1/4 of the standard intravenous thrombolytic dose for patients with acute ischemic stroke due to LVO following successful recanalization.^8^ However, it did not conduct a dose escalation exploratory trial, leaving the ideal dose that balances efficacy and safety unknown.

Tenecteplase, a third-generation thrombolytic drug, has demonstrated superiority over alteplase in improving excellent functional outcomes and reducing disability at 3 months.^19^ There is an important need to determine which dose provides the best balance of efficacy and safety in patients with LVO following successful EVT. Current trials such as POST-TNK^20^ and ANGEL-TNK^21^ are exploring different doses of intra-arterial tenecteplase in patients with acute ischemic stroke due to LVO following successful EVT, using doses of 0.0625 mg/kg, 0.0625 mg/kg, and 0.125 mg/kg, respectively. However, the IA doses in these trials were determined based on a similar fraction of the IV alteplase dose used in CHOICE (one fourth) or were based on trial steering committee recommendation, rather than dose-escalation study. Therefore, this study will provide insight to characterize the optimal dose of adjunctive intra-arterial tenectepalse administration in these patients.

The DATE trial enrolled its first patient on July 27, 2023, and as of August 25, 2024 has enrolled 205 subjects. Full study completion (including collection on 3-month outcomes) is expected by November 2024. When completed, this trial will provide pivotal data allowing assessment of the optimal dose, safety, and efficacy of intra-arterial tenecteplase in patients with acute ischemic stroke due to LVO following successful EVT.

## Data Availability

No applicable

## Funding

The project was supported by the Key Project of Chongqing Science (2023ZDXM025), National Natural Science Foundation of China (No. 82425021), and Health Joint Medical Research Project and the Major Project of Clinical Research Incubation at the First Affiliated Hospital of Army Medical University (2023IITZD01).

## Conflicts of interest

The author(s) declare the following potential conflicts of interest with respect to the research, authorship, and/or publication of this article: JLS reports consulting fees for advising on rigorous and safe clinical trial design and conduct from Biogen, Boehringer Ingelheim, Genentech, Johnson & Johnson, Phenox, Phillips, Rapid Medical, and Roche. TNN discloses Associate Editor of Stroke, advisory board of Aruna Bio and, Brainomix. RGN reports consulting fees for advisory roles with Anaconda, Biogen, Cerenovus, Genentech, Philips, Hybernia, Hyperfine, Imperative Care, Medtronic, Phenox, Philips, Prolong Pharmaceuticals, Stryker Neurovascular, Shanghai Wallaby, Synchron, and stock options for advisory roles with Astrocyte, Brainomix, Cerebrotech, Ceretrieve, Corindus Vascular Robotics, CrestecBio Inc., Euphrates Vascular, Inc., Vesalio, Viz-AI, RapidPulse and Perfuze. RGN is one of the Principal Investigators of ENDOLOW trial. Funding for this project is provided by Cerenovus. RGN is the Principal Investigator of the DUSK trial. Funding for this project is provided by Stryker Neurovascular. RGN is an investor in Viz-AI, Perfuze, Cerebrotech, Reist/Q’Apel Medical, Truvic, Tulavi Therapeutics, Vastrax, Piraeus Medical, Brain4Care, Quantanosis AI, and Viseon. T.Nguyen reports Associate Editor of Stroke; advisory board of Brainomix, Aruna Bio; speaker for Genentech and Kaneka. Other author(s) declare no potential conflicts of interest with respect to the research, authorship, and/or publication of I article.

## Contributorship

ZZ, WZ, XH, DW, JLS, TNN, and RGN designed and conceptualized the study. XH, JH, CG, YH, LW, LC, QL, JW, and MW participated in data collection. XH, YH, JH, and CG wrote the manuscript. All authors critically revised and approved the manuscript.

## Acknowledgement

None.

## Notes

### Clinical Trial

ChiCTR2400080624(www.chictr.org.cn)

## References

1. Global, regional, and national burden of stroke and its risk factors, 1990-2019: a systematic analysis for the Global Burden of Disease Study 2019. The Lancet Neurology. 2021;20(10):795–795.

2. Powers WJ, Rabinstein AA, Ackerson T, et al. Guidelines for the Early Management of Patients With Acute Ischemic Stroke. 2019 Update to th. 2018 Guidelines for the Early Management of Acute Ischemic Stroke: A Guideline for Healthcare Professionals From the American Heart Association/American Stroke Association. Stroke. 2019;50(12):e344–e418.

3. Goyal M, Menon B, van Zwam W, et al. Endovascular thrombectomy after large-vessel ischaemic stroke: a meta-analysis of individual patient data from five randomised trials. Lancet (London, England). 2016;387(10029):1723–1723.

4. Liebeskind DS, Bracard S, Guillemin F, et al. eTICI reperfusion: defining success in endovascular stroke therapy. J Neurointerv Surg. 2019;11(5):433–433.

5. Ng F, Churilov L, Yassi N, et al. Prevalence and Significance of Impaired Microvascular Tissue Reperfusion Despite Macrovascular Angiographic Reperfusion (No-Reflow). Neurology. 2022;98(8):e790–e801.

6. Laredo C, Rodriguez A, Oleaga L, et al. Adjunct Thrombolysis Enhances Brain Reperfusion following Successful Thrombectomy. Ann Neurol. 2022;92(5):860–860.

7. Jia M, Jin F, Li S, et al. No-reflow after stroke reperfusion therapy: An emerging phenomenon to be explored. CNS neuroscience & therapeutics. 2024;30(2):e14631.

8. Renú A, Millán M, San Román L, et al. Effect of Intra-arterial Alteplase vs Placebo Following Successful Thrombectomy on Functional Outcomes in Patients With Large Vessel Occlusion Acute Ischemic Stroke: The CHOICE Randomized Clinical Trial. Jama. 2022;327(9):826–826.

9. Tsivgoulis G, Katsanos AH, Sandset EC, et al. Thrombolysis for acute ischaemic stroke: current status and future perspectives. The Lancet Neurology. 2023;22(5):418–418.

10. Coutts SB, Berge E, Campbell BC, Muir KW, Parsons MW. Tenecteplase for the treatment of acute ischemic stroke: A review of completed and ongoing randomized controlled trials. International journal of stroke : official journal of the International Stroke Society. 2018;13(9):885–885.

11. Campbell BCV, Mitchell PJ, Churilov L, et al. Effect of Intravenous Tenecteplase Dose on Cerebral Reperfusion Before Thrombectomy in Patients With Large Vessel Occlusion Ischemic Stroke: The EXTEND-IA TNK Part 2 Randomized Clinical Trial. Jama. 2020;323(13):1257–1257.

12. Campbell BC, Mitchell PJ, Kleinig TJ, et al. Endovascular therapy for ischemic stroke with perfusion-imaging selection. N Engl J Med. 2015;372(11):1009–1009.

13. Chan AW, Tetzlaff JM, Gøtzsche PC, et al. SPIRI. 2013 explanation and elaboration: guidance for protocols of clinical trials. BMJ (Clinical research ed). 2013;346:e7586.

14. Zhang X, Xie Y, Wang H, et al. Symptomatic Intracranial Hemorrhage After Mechanical Thrombectomy in Chinese Ischemic Stroke Patients: The ASIAN Score. Stroke. 2020;51(9):2690–2690.

15. Huang JH, Su QM, Yang J, et al. Sample sizes in dosage investigational clinical trials: a systematic evaluation. Drug design, development and therapy. 2015;9:305–312.

16. Yuan Y, Hess KR, Hilsenbeck SG, Gilbert MR. Bayesian Optimal Interval Design: A Simple and Well-Performing Design for Phase I Oncology Trials. Clinical cancer research : an official journal of the American Association for Cancer Research. 2016;22(17):4291–4291.

17. Wang Y, Han S, Qin H, et al. Chinese Stroke Association guidelines for clinical management of cerebrovascular disorders: executive summary an. 2019 update of the management of high-risk population. Stroke and vascular neurology. 2020;5(3):270–270.

18. Hernández-Jiménez M, Abad-Santos F, Cotgreave I, et al. Safety and Efficacy of ApTOLL in Patients With Ischemic Stroke Undergoing Endovascular Treatment: A Phase 1/2 Randomized Clinical Trial. JAMA neurology. 2023;80(8):779–779.

19. Palaiodimou L, Katsanos AH, Turc G, et al. Tenecteplase vs Alteplase in Acute Ischemic Stroke Within 4.5 Hours: A Systematic Review and Meta-Analysis of Randomized Trials. Neurology. 2024;103(9):e209903.

20. Jiacheng Huang CG, Jie Yang, Xiaolei Shi, Chang Liu, Jiaxing Song, Fengli Li, Weilin Kong, Shitao Fan, Zhouzhou Peng, Shihai Yang, Jinfu Ma, Xu Xu, Linyu Li, Zhixi Wang, Nizhen Yu, Wenzhe Sun, Chengsong Yue, Xiang Liu, Dahong Yang, Cheng Huang, Duolao Wang, Raul G. Nogueira, Thanh N. Nguyen, Jeffrey L. Saver, Yangmei Chen, Wenjie Zi. Adjunctive Intra-arterial Tenecteplase after Successful Endovascular Thrombectomy in Patients with Large Vessel Occlusion Stroke (POST-TNK): Study Rationale and Design. European Stroke Journal. 2024.

21. Huo X, Luo G, Sun D, et al. Intra-arterial tenecteplase after successful endovascular therapy (ANGEL-TNK): protocol of a multicentre, open-label, blinded end-point, prospective, randomised trial. Stroke and vascular neurology. 2024.

